# Towards a COVID-19 symptom triad: The importance of symptom constellations in the SARS-CoV-2 pandemic

**DOI:** 10.1101/2021.02.01.21250537

**Authors:** Leander Melms, Evelyn Falk, Bernhard Schieffer, Andreas Jerrentrup, Uwe Wagner, Sami Matrood, Jürgen R. Schaefer, Tobias Müller, Martin Hirsch

**Affiliations:** Institute of Artificial Intelligence, Philipps-University Marburg, Marburg, 35033, Germany; Cardiology Department, University Hospital Gießen and Marburg, Marburg, 35033, Germany; Emergency Department, University Hospital Gießen and Marburg, Marburg, 35033, Germany; Department of Gynaecology, University Hospital Gießen and Marburg, Marburg, 35033, Germany; Department of Gastroenterology, Endocrinology, Metabolism and Infectiology, Philipps-University, Marburg, Germany; Centre for undiagnosed and rare diseases, University Hospital Gießen and Marburg, Marburg, 35033, Germany

**Keywords:** computerized questionnaires, self-anamnesis, disease prevention, public health, digital health, mHealth, Crowdsourcing, COVID, COVID-19, SARS-CoV-2, Symptoms, Clinical features, web application, self-assessment

## Abstract

Pandemic scenarios like SARS-Cov-2 require rapid information aggregation. In the age of eHealth and data-driven medicine, publicly available symptom tracking tools offer efficient and scalable means of collecting and analyzing large amounts of data. As a result, information gains can be communicated to front-line providers. We have developed such an application in less than a month and reached more than 500 thousand users within 48 hours. The dataset contains information on basic epidemiological parameters, symptoms, risk factors and details on previous exposure to a COVID-19 patient. Exploratory Data Analysis revealed different symptoms reported by users with confirmed contacts vs. no confirmed contacts. The symptom combination of anosmia, cough and fatigue was the most important feature to differentiate the groups, while single symptoms such as anosmia, cough or fatigue alone were not sufficient. A linear regression model from the literature using the same symptom combination as features was applied on all data. Predictions matched the regional distribution of confirmed cases closely across Germany, while also indicating that the number of cases in northern federal states might be higher than officially reported. In conclusion, we report that symptom combinations anosmia, fatigue and cough are most likely to indicate an acute SARS-CoV-2 infection.

## Introduction

In December 2019 cases of pneumonia of unknown etiology were reported in Wuhan, China^1^. The pathogenic agent, which was later identified as the severe acute respiratory syndrome coronavirus 2 (SARS-CoV-2) rapidly spread over the whole country and the rest of the world. On January 31, the World Health Organization held a press conference and declared the outbreak a Public Health Emergency of International Concern (PHEIC) ^2^. On March 11, 2020, the coronavirus outbreak was declared a global pandemic by the WHO ^2^. Lockdown measures were introduced on March 22st in Germany ^3^. Up to July 30, there has been more than 16 million confirmed cases worldwide.

The unprecedented speed and infectivity of such a pandemic requires novel information and communication structures for consolidating scientific knowledge ^4–6^. So-called data crowdsourcing has been used as a warning and information system for waves of influenza since late 2000 7–9. This usually refers to applications that query flu-like symptoms at regular time intervals and issue warnings for affected areas by consolidating and analyzing the data. The advantage of these solutions lies in their cost-effective scalability and the resulting speed of data collection ^9,10^, making them a crucial tool for the data-driven response of highly infectious diseases ^11^. With the SARS-CoV-2 outbreak, many of these apps have been adapted or newly developed accordingly: In Switzerland “covidtracker.ch” ^12^, in Spain “CoronaMadrid” ^13^, in France “maladiecoronavirus.fr” ^14^ and in England the App “COVID Symptom Study” ^15^ are successfully used.

Currently, health care authorities primarily monitor the spreading dynamics of SARS-CoV-2 by the number of virus-specific positive reverse transcriptase polymerase chain reaction (RT-PCR) tests. However, the rapid transmission of SARS-CoV-2 has revealed weaknesses in current semi-digital monitoring practices of federally organized states like Germany, which heavily rely on decentralized reporting of laboratory tests. For an effective early warning system, however, daily updated data streams are of essential importance in order to be able to take effective measures quickly. Analysis of crowd-sourced clinical features and risk factors offers a promising opportunity for continuous prevalence monitoring in the population. In the future, both monitoring methods could perfectly complement each other in order to better understand the dynamics of infectious transmissions ^6^.

Numerous studies have shown that the SARS-CoV-2-infection has diverse signs and symptoms ^16–19^, with fever, cough and fatigue being the most common at onset of illness ^19–23^. However, recent studies suggest that only half the patients are febrile at the time of hospital admission ^19,22^. In addition, more attention is now drawn to formerly underestimated extrapulmonary symptoms and symptom constellations ^24–28^: loss of smell (i.e. anosmia), for instance, varies greatly in COVID-19 patient (33 – 68%) ^29^ but often was one of the first apparent symptoms ^30^. In Germany, the Robert Koch Institute (RKI) responsible for disease control and prevention, records loss of smell and taste as symptoms for COVID-19 cases since calendar week 17. According to current RKI recommendations, patients are now also being tested for SARS-CoV-2, who show impairments of the sense of smell ^31^. Already in March, the British Society of ENT Physicians, ENT UK, wrote in a statement: “There is already good evidence from South Korea, China and Italy that significant numbers of patients with proven COVID-19 infection have developed anosmia/hyposmia” ^32^. The British physicians called for people who noticed a loss of smell to voluntarily quarantine themselves. The American Academy of Otolaryngology-Head and Neck Surgery (AAO-HNS) also reported that a loss of smell has been observed in otherwise symptom-free SARS-CoV-2 positives and, as a consequence, developed a COVID-19 Anosmia Reporting Tool ^33^. In a collaborative effort, the COPE consortium of UK- and US-based researches investigated a smartphone-based self-reported longitudinal and diverse COVID-19 survey data set (n = 805753) and found that the less common symptom anosmia in combination with the more established symptoms fever and cough were the strongest predictors of COVID-19 ^24^, complimenting recent scientific findings.

## Methods

### Technology and study participants

The symptom checking application COVID-Online was developed by the Institute of Artificial Intelligence (Philipps University Marburg) and is written in the Go programming language. The web app was accessible via a publicly accessible website covid-online.de. The questionnaire contained a total of 38 items, whereby 10 items appear only in relation to previous items (i.e. adaptive items).

Informed consent for scientific evaluation was provided by accepting the privacy policy (explicitly asked before submitting the questionnaire, see point 3.1 in the privacy policy). Users also had to confirm that they are at least 18 years old. Furthermore, the paragraph “I agree that COVID-Online may process and evaluate the data I have entered in a pseudonymised form” had to be explicitly agreed to as an opt-in checkbox.

The graphical user interface to answer the questions contained single choice, multiple choice and free text input fields. The questionnaire was divided into three segments, which were presented one after the other: first, basic epidemiological data such as gender, approximate age, approximate height, and approximate weight were collected. Then current symptoms (fever, body aches, cough, sniff, diarrhea, nausea, vomiting, throat pain, headache, loss of smell or taste, dyspnea, fatigue) were queried that could indicate COVID-19. Finally, users were prompted for individual risk factors such as smoking status and comorbidities (lung disease, diabetes, cardiovascular disease, stroke, tumor disease, chronic inflammatory disease, kidney disease, allergies), vaccination status (flu, measles) and their postal code. The complete survey items are available in the supplementary (Table 1).

**Table 1:** Base characteristics of the study population.

| Variable | n | % |
| --- | --- | --- |
| Age |  |  |
| - < 20 | 56676 | 08·42 |
| - 20 – 30 | 144652 | 21·49 |
| - 31 – 40 | 210658 | 31·29 |
| - 41 – 50 | 126731 | 18·82 |
| - 51 – 60 | 80993 | 12·03 |
| - 61 – 70 | 36801 | 05·47 |
| - > 70 | 16647 | 02·47 |
| Gender |  |  |
| - Male | 392326 | 58·28 |
| - Female | 279642 | 41·54 |
| - Other | 1190 | 00·17 |
| Weight |  |  |
| - < 60kg | 70690 | 10·50 |
| - 60 – 70 kg | 115590 | 17·17 |
| - 71 – 80 kg | 134079 | 19·91 |
| - 81 – 90 kg | 135229 | 20·09 |
| - 91 – 100 kg | 96334 | 14·31 |
| - 101 – 110 kg | 57436 | 08·53 |
| - 111 – 120 kg | 32141 | 04·77 |
| - > 120 kg | 31659 | 04·70 |
| Height |  |  |
| - < 1.50 m | 6036 | 00·90 |
| - 1.50 – 1.60 m | 52106 | 07·74 |
| - 1.61 – 1.70 m | 178958 | 26·58 |
| - 1.71 – 1.80 m | 235466 | 34·98 |
| - 1.81 – 1.90 m | 165267 | 24·55 |
| - 1.91 – 2.00 m | 33193 | 04·93 |
| - > 2.00 m | 2132 | 00·32 |

The web app was launched on April 3rd. At this time, the new infections reached its peak in Germany and the population was in a phase of great uncertainty. Our main goal developing COVID-Online was patient care navigation by guiding the participant through the next steps in case of increased risk, and by this serving as a guidance system for regional patient management to enable an efficient allocation of resources in case of emergency.

## Data analysis

### Data preparation and data management

All data was obtained from a database query (SQL) in the comma-separated values format (CSV) from the application servers. Program libraries Pandas, SciPy, Matplotlib, Seaborn and Jupyter were used for exploratory data analysis, data post-processing and plotting. We dropped users with missing or invalid postal codes.

### Statistical evaluation

Basic characteristics were analysed using descriptive statistics. Groups were formed based on the independent variables age, gender, comorbidities and confirmed contact to a COVID-19 patient. Chi-squared test and Cramer’s V correlation were used for data analysis. For categorical variables, we report absolute numbers and percentages. For continuous variables, we report averages ± standard deviation, unless otherwise stated. No imputation was made for missing data. All statistical tests were carried out with a significance level of α = 5%.

### Hypothesis testing

Previous work by Drew et al. has shown that symptom combinations are more suitable for distinguishing between COVID-19 positive and negative cases than single symptoms alone ^34^. We investigated if the same applies to our dataset. Exploratory data analysis revealed that groups can be divided based on the question “I had confirmed contact to a COVID-19 case”. The reason for a different reporting of symptoms of subjects with confirmed contact could be that these participants have a higher sensitivity to symptom inquiry due to fear of infection or that contact has led to a manifestation of the disease with corresponding presentation of symptoms. In the latter case, a higher overall level of infection is to be expected within the formed group. Lastly, we applied the generated linear model by Drew et al. including age, sex, loss of smell, fatigue, cough and loss of appetite to our data:

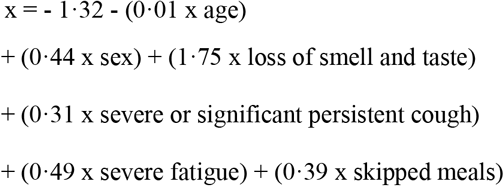

where the prediction was calculated as exp(x) / (1 + exp(x) with a threshold of 0·5 as suggested by Drew et al. ^34^. As loss of appetite was not queried by our questionnaire, it was set to true whenever a subject reported anosmia. Clinical experience has shown that loss of appetite rarely occurs isolated and is rather a consequence of the loss of smell. To confirm our findings, we compared the frequencies of predicted and confirmed infections in a seven-day window geographically by districts and contrasted this with the distribution of the individual predictive symptoms fever, anosmia and cough.

## Results

A total of 712,018 users completed and submitted the survey from April 3 to July 1. After post-processing and deletion of false or incomplete data entries 673,158 submissions remained. Baseline characteristics are summarized in table 1. The approximate age was recorded with a single choice item and included the groups < 20 years old (8·42 %, *n* = 56676), 20 - 30 years old (21·49 %, *n* = 144652), 31 - 40 years old (31·29 %, *n* = 210658), 41 - 50 years old (18·83 %, *n* = 126731), 51 - 60 years old (12·03 %, *n* = 80993), 61 - 70 years old (5·47 %, *n* = 36801) and over 70 years old (2·47 %, *n* = 16647). The subjects were 58·28 % (n = 392326) male, 41·54 % (*n* = 279642) female and 0·18% (*n* = 1190) other.

Exploratory Data Analysis revealed that the answer to the question whether if a previous contact with a confirmed case of a COVID patient occurred, changed the symptom frequency distribution between these groups.

A chi-square test of independence was performed to examine the relation between previous confirmed contact and individual symptoms sniff, cough, fatigue, body aches, headache, diarrhea, sore throat, nausea, dyspnea at rest, anosmia and fever. The relation between these variables was significant (Table 2):

**Table 2:** Results of the chi-square test of independence

| Variable | X2 | p-value | Degrees of freedom |
| --- | --- | --- | --- |
| sniff | 551.734708 | < .001 | 1 |
| cough | 1671.296687 | < .001 | 1 |
| fatigue | 1780.535304 | < .001 | 1 |
| body aches | 2190.363285 | < .001 | 1 |
| headache | 2232.331668 | < .001 | 1 |
| diarrhea | 2389.594631 | < .001 | 1 |
| sore throat | 2630.390694 | < .001 | 1 |
| nausea | 4716.380914 | < .001 | 1 |
| dyspnea at rest | 5297.727920 | < .001 | 1 |
| anosmia | 7484.169351 | < .001 | 1 |
| fever | 8779.939422 | < .001 | 1 |

Therefore, patients were divided bases on the dichotomous question “I had contact to a confirmed case of COVID 19” into two groups.

Figure 1 illustrates the importance of individual symptoms in distinguishing between the two groups (Chart A) on the one hand and, as a result, indicates that only a combination of several symptoms can achieve sufficient differentiation between the groups on the other hand (Chart B). Interestingly, the frequencies of single symptoms dominate in both groups, with fatigue, cough and diarrhea being the most common. However, in the group with confirmed contact with a COVID-19 patient, not only is anosmia more common as a single symptom, but also the frequency of triple combinations of symptoms in comparison. In the group without confirmed contact these triple combinations are much less frequent. Also, the frequencies of symptoms drop much faster in the group without confirmed contact than the frequencies of symptom combinations within the confirmed contact group. This confirms the impression that single symptoms and symptom combinations less or equal than two might be less suitable for the (geographical) identification of COVID-19 patients than the combination of more symptoms, whereby the combination of fatigue, cough and anosmia is particularly discriminative. This particular symptom combination, which combines the more frequent symptoms cough and fatigue with the less frequent symptom of anosmia, was also the most predictive in the work by Drew et al., upon which their scoring for COVID-19 was build on. Furthermore, the model of Drew et al. predicts a percentage of 23·21% (*n* = 4439) positive cases within the confirmed contact group (*n* = 19128) and only 6·21% (*n* = 1233) positive cases within a equally-sized random sample from our poplation without confirmed contact. This suggests – taking into account the work of Drew et al. – that in the group of confirmed contacts more positive cases must exist.

**Fig 1:**
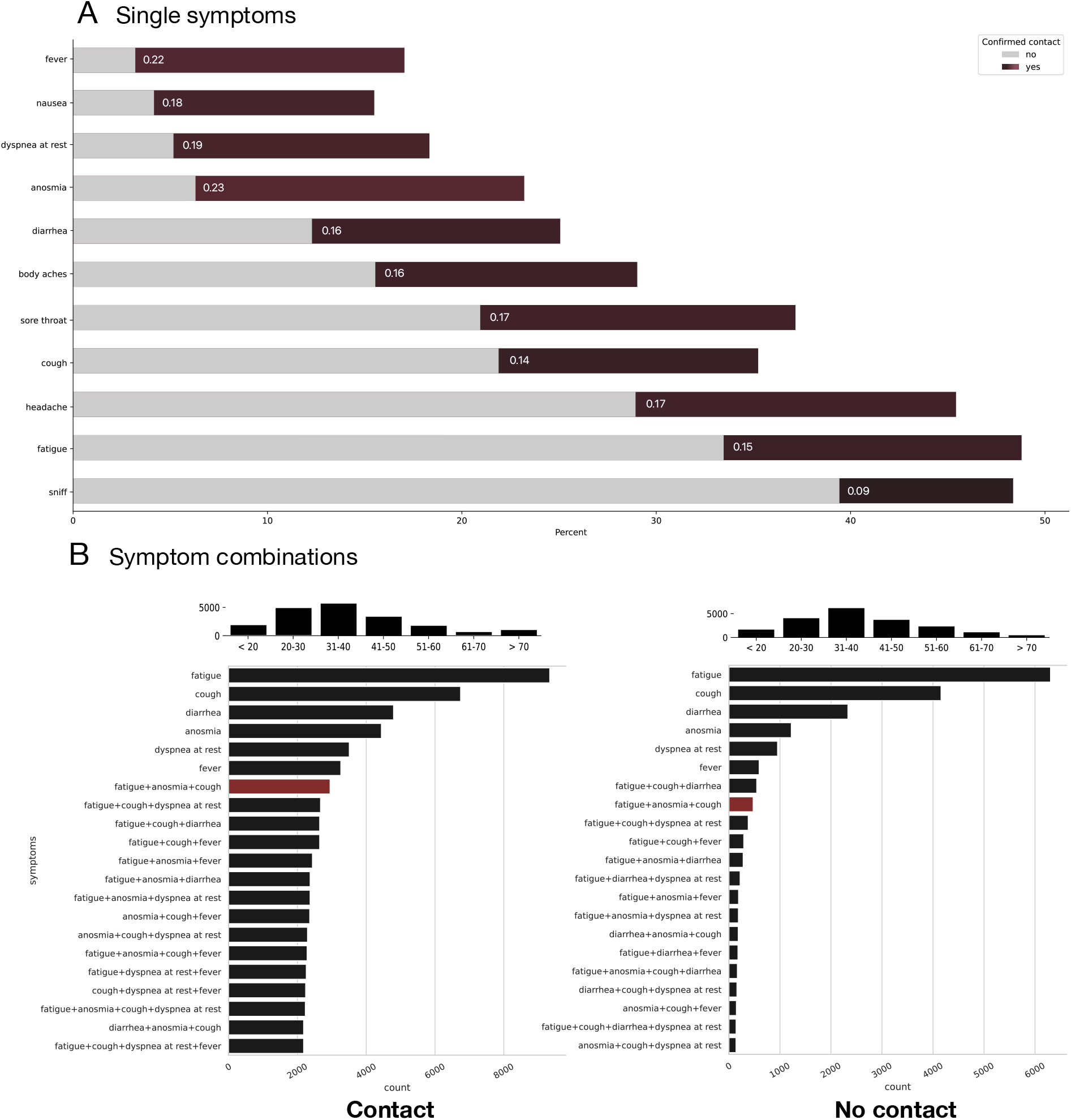
Comparison of symptom distribution between patients with and without confirmed contact. **A) Single symptoms**: Color coded Cramer’s V correlation of symptoms with the confirmed contact variable (dark red tones) & symptom frequency count in percent of positive statements broken down by groups with and without confirmed contact. Anosmia seems to be the strongest predictor followed by fever and dyspnea at rest. On the contrary, the least single important symptoms are sniff, fatigue and cough by itself. **B) Complex symptoms**: Symptom frequency count (total) with combinations. The symptom combination fatigue, anosmia and cough has been highlighted in red to illustrate the shift of importance between the two groups. The age distribution of the two groups is depicted in each case above graph B. A random sample of 19128 was taken from the population without confirmed contact for comparison. The percentage of positive cases in the total number of participants without contact was 6.24% whereas the percentage of positive cases in the group with confirmed contact was 23.21%.

Figure 2 shows six maps (A – F) of Germany: In the upper row, map A displays the frequency of participants per 100 thousand inhabitants per district in a colour-correlated manner. Map B depicts the number of predicted infections based on the model by Drew et al. and map C the number of confirmed infections per 100 thousand inhabitants. Maximum value of 50 infections per 100 thousand inhabitants per district in the last 7 days was chosen because of the respective lockdown regulations in effect in Germany.

**Fig 2:**
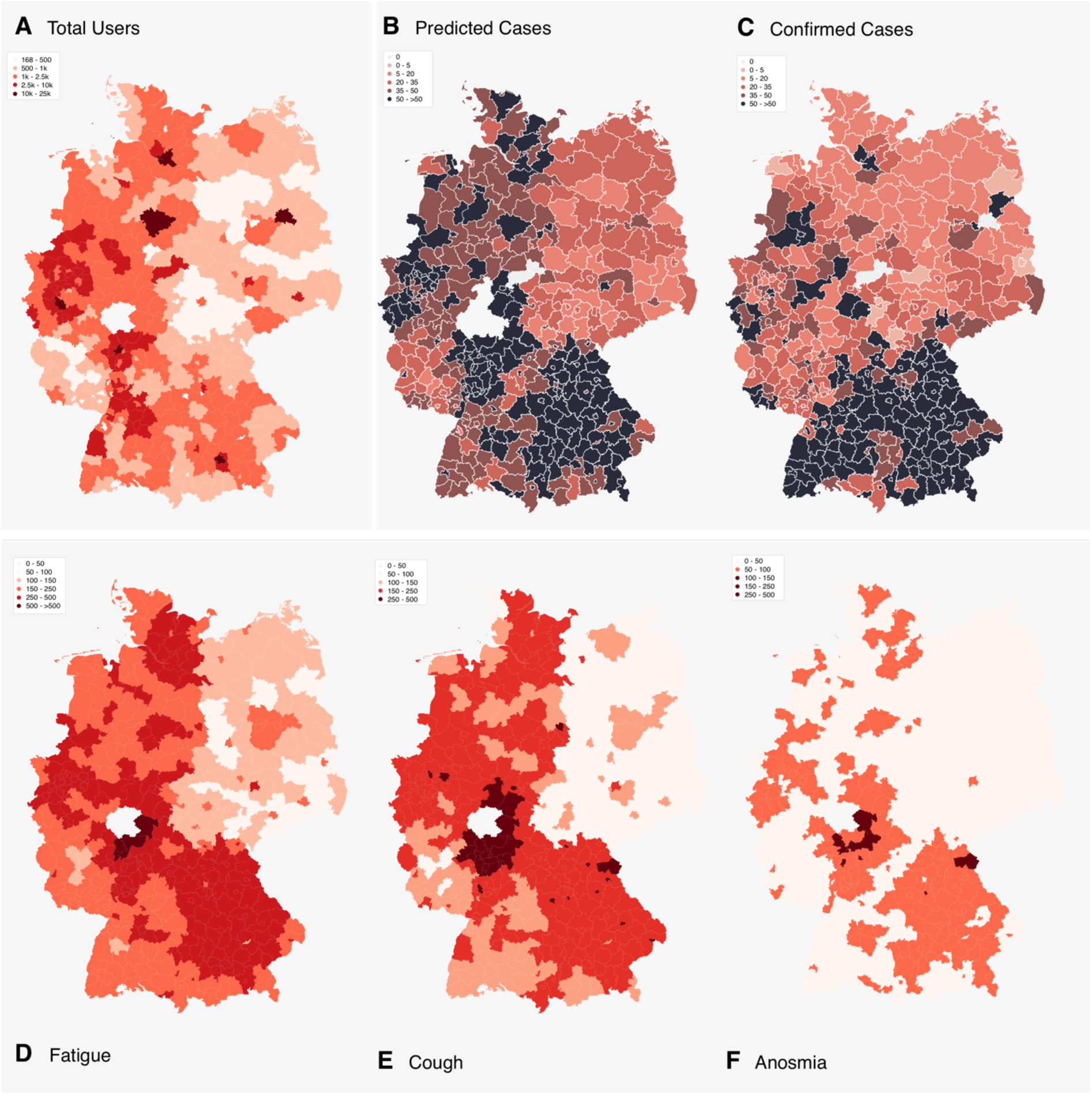
Maps illustrating the total number of users of COVID-Online (A), the number of predicted infections (B), the number of confirmed infections (C), the frequency of fatigue (D), the frequency of anosmia (E) and the frequency of fever (F). Data of maps A – F is based on the time period 03.04 – 10.04.2020. **A, B, D, E, F**: The district “Marburg-Biedenkopf” has been excluded from these charts as it contained too many records from internal tests carried out by associated personnel of COVID-Online and was also influenced by regional media reports. Unfortunately, due to the great time pressure in times of crisis, no test or staging instance could be installed for such purposes.

The comparison between the map with the frequency of all users per 100 thousand inhabitants per district with the map with the frequency of predicted infections reveals that the frequencies of predicted infections are largely independent of the number of users in the district. This is an important indicator for the specificity of the applied scoring model. Furthermore, the frequencies of individual symptoms (maps D - F) cannot be clearly assigned to the confirmed infections in comparison to the map of predicted infections using complex symptoms, again confirming that only combinations of symptoms are sufficiently discriminatory.

When comparing the predicted infections with the confirmed infections (Maps B and C), it can first of all be noted that the most overlaps are found in the most severely affected states of Bavaria and Baden-Wuerttemberg. There are also isolated overlaps in North Rhine-Westphalia and Lower Saxony. According to the predicted infections, the northern part of Germany, and in particular Schleswig-Holstein, are much more severely affected than the number of confirmed cases suggest. This may be due to insufficient specificity of the score, increased multiple participation, incorrect reporting in northern Germany and/or insufficiently performed tests in northern Germany. Another reason for differences between the distribution of confirmed infections and predicted infections is that the duration of symptoms of confirmed positive persons can last 2 - 3 weeks even in non-hospitalized patients ^35^. Anosmia regresses significantly later in some patients. Tenforde et al found that it takes from onset to clinical recovery about 2 weeks for mild cases and 3-6 weeks for patients with severe or critical disease; the symptoms to least likely have resolved within 14–21 days after the test date included and fatigue ^35^. This means that people who tested positive 2 - 3 weeks ago do not show up in the statistics of the last 7 days, but they still report their symptoms in a symptom checker and appear in the statistics of predicted infections. On the contrary, confirmed cases do not necessarily need to show symptoms, as symptoms could have been resolved before the test date or started after the date of testing.

Unfortunately, there is not enough data on the number of total tests, because in Germany, according to the Infection Protection Act, only positive cases are reported to the health authorities. This makes it difficult to determine the actual test density per federal state and even more so per district. However, laboratories are invited to send the number of tests to the RKI. Since participation is voluntary, no statement can be made about the completeness of the tests (see Table 3 and 4):

**Table 3:** Number of tests submitted by 177 laboratories voluntarily in calendar week 14 (06.04 – 12.04.2020, source: RKI). Data is from a sample of laboratories, not a complete survey of all tests in Germany. The coverage and representativeness of the data can greatly vary between the federal states. Unfortunately, the proportion of voluntary participation of the total is not known. Up to and including week 14/2020, 177 laboratories have registered for this RKI test laboratory interrogation or in one of the other transmitting networks. The data was derived from the Management Report on Coronavirus Disease (as of April 8, 2020) and the graphs contained therein.

| Federal state | N tests / calendar week 15 2020 | Tests / 100k residents |
| --- | --- | --- |
| BW | ~ 5 000 | ~ 45·17 – 67·75 |
| BY | ~ 15 000 | ~ 114·29 |
| NI | ~ 5 000 | ~ 62·55 |
| SH | ~ 1 500 – 2 500 | ~ 51·78 – 86·30 |
| NRW | ~ 25 000 | ~ 139·30 |

**Table 4:** Number of SARS-CoV-2-PCR tests (cumulative) broken down by federal state. Data status until 23.04.2020 (source RKI). The percentage of the total tests to the number of inhabitants of a federal state was displayed in green from a value of 0.5 and otherwise in red. Number of conducted tests greatly varied between federal states. Populous federal states in the south and west of Germany have performed significantly more tests than the northern and eastern regions.

| Federal state | N | N positive | % positive | Population | % tested |
| --- | --- | --- | --- | --- | --- |
| NRW | 148,968 | 12,128 | 8·1 | 17,947,221 | 0·8 |
| BY | 91,463 | 9,480 | 10·4 | 13,124,737 | 0·7 |
| BW | 46,265 | 5,514 | 11·9 | 11,069,533 | 0·4 |
| NI | 39,563 | 1,893 | 4·8 | 7,993,608 | 0·5 |
| RP | 35,784 | 2,626 | 7·3 | 4,093,903 | 0·9 |
| SH | 7,860 | 313 | 4 | 2,896,712 | 0·3 |
| MV | 4,389 | 100 | 2·3 | 1,608,138 | 0·3 |

From these tables it can be seen that Bavaria and North Rhine-Westphalia have tested the most, both in terms of population and in absolute terms. Lower Saxony, Baden-Wuerttemberg, Schleswig-Holstein and Mecklenburg-Western Pomerania, on the other hand, tested least according to these data. However, it must be emphasised that this may also be due to a lower participation rate in data transmissions. If one assumes, however, that the participation density between the federal states would be approximately the same, more cases would probably have been measured in these states.

In contrast to other countries, preventive testing is not carried out in Germany to date, but only as an indication test on symptomatic patients and contact persons of such patients. Furthermore, the RKI recommends extending the indication for testing to nursing or care facilities and medical personnel. In the case of local outbreaks, mass testing is also carried out within a few days to contain the outbreak.

## Discussion

The study investigated whether symptoms or symptom constellations are indicative of COVID-19. Moreover, it was examined to what extent a model, that accept such symptom constellations as input features, can accurately estimate the number of positive cases.

Previous studies have shown that symptom combinations of equal to or greater than three are more indicative of COVID-19 while predictions based on single symptoms such as anosmia, coughing or fatigue alone are less specific ^24,34^. This corresponds with the results of this work. The triple combination of anosmia, fever and cough was most important in distinguishing between groups of participants with and without active contact. The model published by Drew et al. generated 23.21% positive cases in the group with confirmed contact whereas the percentage of positive cases in the group without confirmed contact was 6.24%. This supports the assumption that there must be more COVID-positive patients in the group with confirmed contact. It is moreover remarkable that the same combination of symptoms of anosmia, fever and cough could be determined as the most important feature in this work al by Drew et al. The analysis of the distribution of the predicted positive cases among the districts showed that the distribution of the confirmed positive cases largely overlapped, especially in the federal states of Bavaria, Baden-Wuerttemberg and North Rhine-Westphalia. If the density of testing in the federal states is also considered, it can be assumed that the number of positive cases in the northern part of Germany is likely to be higher at the time of observation. In these cases, more test resources could be mobilized to initiate effective isolation and containment strategies for the spread in the future.

The near real-time analysis of symptom tracking applications might help to more accurately understand the spread and infectivity of infectious diseases such as SARS-CoV-2. In addition, risk factors can be collected and quantified in addition to symptoms. This is a decisive added value compared to the sole laboratory testing for an infectious disease, since persons at risk can be identified on the basis of the data and can be more quickly assigned to a potential therapy. The user numbers from Germany and England indicate a broad acceptance of such tools in pandemic scenarios ^24,34^. Symptom-tracking tools may thus play an important role in the control and containment of infectious diseases, alongside Sars-CoV-2, and provide significant added value for public health matters. Future implications are based on these tests and their accuracies, that the combination of both, adequate laboratory tests plus preventive screening such as COVID-Online, may predict future outbreaks and/or hint towards hotspot developments in clinically actionable windows.

It must be emphasized, though, that symptom screening and thereby symptom-based scoring systems will not likely reach a sufficient sensitivity for COVID-19 as asymptomatic carriage remains a problem. For instance, in the Diamond Princess cruise ship study, it was estimated that a proportion of 17.9% among all infected cases were asymptomatic ^36^. Asymptomatic patients further complicate the screening problem by the high risk of silent transmission. This implicates that the value of symptom inquiry for individual screening scenarios might be limited, but nevertheless offers crucial insights on a public health level when it comes to understanding the dynamics of spread, containment strategies and hot spot identification.

### Limitations of the study

Our work has numerous limitations. One major limitation of our study is that we do not have data on confirmatory laboratory test results of the study participants. Because of this, we cannot extend and/or adopt the proposed model by Drew et al. by means of, for instance machine learning. Notably though, the same symptom combination was the most discriminative between the groups with and without confirmed contact, which suggests that the proposed combination might indeed achieve the highest specifity. In addition, we recognize that the data collected originates from a modern web application. Although a balanced gender and age distribution was achieved in the study population, it can be assumed that less technology-affine individuals are underrepresented in the study collective, which means that a contributor bias cannot be excluded. Thus, it cannot be ruled out that certain symptom constellations are more likely to be significant than others, which may be more common in underrepresented user groups. Furthermore, the nature of the collected data is self-reported. Data validity is therefore not checked and may contain false statements. Another limitation is that since users cannot create an account for reasons of data security and privacy, only snapshots of symptoms can be collected and cannot be tracked over time (i.e. longitudinal data). Also, our data and calculations are based on submissions of the questionnaire and do not strictly correlate with the number of individuals, as users can access the web app and submit the questionnaire multiple times. Because of this, we are only able to identify potential COVID-19 patients at the time of complete manifestation.

## Conclusions

With the enormous advancements in technology we now have valuable digital tools that not only can keep up with the speed of the spread but also offer unique and actionable key insights for health care professionals. Our work confirms that the symptom combination of anosmia, fatigue and cough are indeed more specific for COVID-19 than single symptoms alone. Besides identifying potential hot spots that require more test resources, data on pre-existing conditions and risk factors such as smoking are obtained, too. By identifying high risk patients with a high probability of a SARS-CoV-2 infection, this opens up the opportunity to take timely preventive measures. While asymptomatic carriage remains an issue, our work supports the recommendation that patients who are suffering from anosmia, fever and cough should consider self-quarantine. Moreover, our approach demonstrates that crowd-sourced data represents an effective and scalable means for collecting population-based data for a data-driven public health response to infectious disease outbreaks in light of the COVID-19 pandemic. We could show that sophisticated and well-accepted real-time symptom reporting analytical platforms can be developed rapidly, even with limited personnel resources. We also demonstrate the potential of crowdsourced data to complement traditional public health surveillance methods, primarily through providing fast, on-demand insights and increased information coverage.

## Data Availability

The survey data analyzed in study is not publicly available due to information that could compromise patient privacy/consent.

## Acknowledgements

The authors thank all the participants, coordinators, and administrators for their support and help during the data acquisition. This work has received financial support from the Münch Foundation. The founding source did not have any additional role in the study design, data collection and analysis, decision to publish, or preparation of the manuscript. Our thanks for active medical support go especially to: Dr. med. Wollenberg, Dr. med. Yulia Sharkova, Prof. Dr. med. Jürgen Schäfer, Prof. Dr. med. Uwe Wagner, Prof. Dr. Harald Renz, Markus Müller, Maik Klein Dr. med. Hartmut Hesse and PD Dr. med. Timm Greulich.

## Informed Consent

The study was reviewed by the medical ethics committee of the Philipps-University Marburg (reference number: 59/20). All research was performed in accordance with current guidelines and regulations.

## Author contributions

L.M., E.F., B.S., M.H., A.J. and J.R.S. conceived and designed the study. L.M. collected, reviewed and analysed the data. S.M. helped in data collection. T.M. reviewed and analysed the data. L.M. wrote and coded the application used in this study. L.M. and E.F. wrote the manuscript and B.S, M.H. J.R.S., A.J., U.W, S.M, T.M. reviewed it. All authors read and approved the final manuscript. All authors had full access to all the data.

## Additional information

All correspondences and requests for materials should be addressed to L.M.

