## Supplemental Table 1 for "Towards a COVID-19 symptom triad: The importance of symptom constellations in the SARS-CoV-2 pandemic"

[*] Leander.Melms [at] uk-gm [dot] de

**Table 1: Survey questions of the COVID-Online application, divided by languages (German, English, French).**

| **Allgemeine Angaben** | **General information** | **Informations générales** |
|---|---|---|
| 1. Geschlecht | 1. Gender | 1. Sexe |
| a. Weiblich<br>b. Männlich<br>c. Divers | a. Female<br>b. Male<br>c. Diverse | a. Féminin<br>b. Masculin<br>c. Divers |
| 2. Größe | 2. Height | 2. Taille |
| a. Kleiner als 1,50 m<br>b. 1,51 bis 1,60 m<br>c. 1,61 bis 1,70 m<br>d. 1,71 bis 1,80 m<br>e. 1,81 bis 1,90 m<br>f. 1,91 bis 2,00 m<br>g. Über 2,00 m | a. < 1.50 m<br>b. 1.51-1.60 m<br>c. 1.61-1.70 m<br>d. 1.71-1.80 m<br>e. 1.81-1.90 m<br>f. 1.91-2.00 m<br>g. > 2.00 m | a. < 1.50 m<br>b. 1.51-1.60 m<br>c. 1.61-1.70 m<br>d. 1.71-1.80 m<br>e. 1.81-1.90 m<br>f. 1.91-2.00 m<br>g. > 2.00 m |
| 3. Gewicht | 3. Body weight | 3. Poids |
| a. Weniger als 60kg<br>b. 61-70 kg<br>c. 71-80 kg<br>d. 81-90 kg<br>e. 91-100 kg | a. < 60 kg<br>b. 61-70 kg<br>c. 71-80 kg<br>d. 81-90 kg<br>e. 91-100 kg | a. < 60 kg<br>b. 61-70 kg<br>c. 71-80 kg<br>d. 81-90 kg<br>e. 91-100 kg |

| | | |
|---|---|---|
| f. 101-110 kg<br>g. 111-120kg<br>h. Mehr als 120kg | f. 101-110 kg<br>g. 111-120 kg<br>h. > 120 kg | f. 101-110 kg<br>g. 111-120 kg<br>h. > 120 kg |
| 4. Wie alt sind Sie? | 4. How old are you? | 4. Quel age avez-vous? |
| a. Jünger als 20<br>b. 20-30<br>c. 31-40<br>d. 41-50<br>e. 51-60<br>f. 61-70<br>g. 71-80<br>h. Über 80 | a. < 20<br>b. 20-30<br>c. 31-40<br>d. 41-50<br>e. 51-60<br>f. 61-70<br>g. 71-80<br>h. > 80 | a. < 20<br>b. 20-30<br>c. 31-40<br>d. 41-50<br>e. 51-60<br>f. 61-70<br>g. 71-80<br>h. > 80 |
| **Krankheitsspezifische Fragen**<br>Scoring für Wahrscheinlichkeit einer Covid-19-Infektion | **Disease-specific questions**<br>Scoring for the probability of Covid-19 infection | **Questions spécifiques à la maladie**<br>Score pour la probabilité d'infection Covid-19 |
| 5. Hatten Sie engen Kontakt zu einem bestätigten Coronafall? | 5. Did you have close contact with a confirmed Corona case? | 5. Étiez-vous en contact étroit avec un cas Corona confirmé? |
| **a.** Ja → Wann war der letzte Kontakt? Anmerkung: Bitte nehmen Sie Kontakt zur Hotline des regionalen Gesundheitsamtes auf. Gesundheitsamt Marburg: Tel.: 06421-40544<br>1. innerhalb der letzten 14 Tage<br>2. länger als 14 Tage her<br><br>**b**. Nein → Hatten Sie engen Kontakt zu einem Verdachtsfall?<br>1. Ja → Wann war der letzte Kontakt?<br>i. innerhalb der letzten 14 Tage<br>ii. länger als 14 Tage her<br>2. Nein | **a.** Yes → When was the most recent contact? Note: Please contact the hotline of your local health department. Health Department of Marburg: Phone +49 (0) 6421-40544<br>1. within the last 15 days<br>2. more than 15 days ago<br><br>**b.** No → Did you have close contact with a suspected Corona case?<br>1. Yes → When was the most recent contact?<br>i. within the last 15 days<br>ii. more than 15 days ago<br>2. No | a. Oui → À quand remonte le dernier contact? Remarque: veuillez contacter la hotline de votre service de santé local. Département de la Santé de Marburg: Tel.: +49 (0)6421-40544<br>1. au cours des derniers 15 jours<br>2. il y a plus de 15 jours<br><br>b. Non → Étiez-vous en contact étroit avec un cas suspect de Corona?<br>1. Oui → à quand remonte le dernier contact?<br>i. au cours des dernier 15 jours<br>ii. il y a plus de 15 jours<br>2. Non |
| 6. Haben Sie oder hatten Sie Fieber über 38,5°C? (in den letzten 5 Tagen) | 6. Do or did you have a fever above 38.5°C? (within the last 5 days) | 6. Avez-vous ou aviez-vous une fièvre supérieure à 38,5°C? (au cours des 5 derniers jours) |
| a. Ja<br>b. Nein | a. Yes<br>b. No | a. Oui<br>b. Non |
| 7. Haben Sie Gliederschmerzen | 7. Do you have body aches? | 7. Avez-vous des courbatures? |
| a. Ja<br>b. Nein | a. Yes<br>b. No | a. Oui<br>b. Non |
| 8. Haben Sie anhaltenden Husten? | 8. Do you have persistent cough? | 8. Avez-vous une toux persistante? |

| | | |
|---|---|---|
| a. Ja → Haben Sie trockenen Husten oder haben Sie Husten mit Auswurf (Schleim)?<br>1. Trockener Husten<br>2. Husten mit Auswurf<br>b. Nein | a. Yes → Do you have dry cough or do you have cough with expectoration (mucous)?<br>1. Dry cough<br>2. Cough with expectoration<br>b. No | a. Oui → Avez-vous une toux sèche ou avez-vous une toux avec flegme?<br>1. Toux sèche<br>2. Toux avec flegme<br>b. Non |
| 9. Haben Sie eine laufende oder verstopfte Nase? (Schnupfen) | 9. Do you have symptoms of common cold? | 9. Avez-vous des symptômes de rhume? |
| a. Ja<br>b. Nein | a. Yes<br>b. No | a. Oui<br>b. Non |
| 10. Haben Sie Durchfall? | 10. Do you suffer from diarrhea? | 10. Avez-vous de la diarrhée? |
| a. Ja<br>b. Nein | a. Yes<br>b. No | a. Oui<br>b. Non |
| 11. Haben Sie Übelkeit und Erbrechen? | 11. Do you suffer from nausea or vomiting? | 11. Avez-vous des nausées et des vomissements? |
| a. Ja<br>b. Nein | a. Yes<br>b. No | a. Oui<br>b. Non |
| 12. Haben Sie Halsschmerzen? | 12. Do you have a sore throat? | 12. Avez-vous un mal de gorge? |
| a. Ja<br>b. Nein | a. Yes<br>b. No | a. Oui<br>b. Non |
| 13. Haben Sie Kopfschmerzen? | 13. Do you have headaches? | 13. Avez-vous mal à la tête? |
| a. Ja<br>b. Nein | a. Yes<br>b. No | a. Oui<br>b. Non |
| 14. Haben Sie Veränderungen der Geschmacks- oder Geruchswahrnehmung bemerkt? | 14. Did you notice changes in taste or smell? | 14. Avez-vous remarqué des changements de goût ou d'odeur? |
| a. Ja<br>b. Nein | a. Yes<br>b. No | a. Oui<br>b. Non |
| 15. Haben Sie deutliche Luftnot in Ruhe? (Luftnot: Sprechen eines Satzes ist nur mit zusätzlichen Atemzügen möglich) | 15. Do you have shortness of breath at rest? (Shortness of breath: speaking a sentence is only possible with additional breaths) | 15. Avez-vous un essoufflement au repos? (Essoufflement: prononcer une phrase n'est possible qu'avec des respirations supplémentaires) |
| a. Ja → Empfinden Sie die Luftnot als bedrohlich?<br>1. Ja<br>2. Nein<br>b. Nein → Kommen Sie außer Atem, wenn Sie mehr als 30 Meter gehen oder 10 Treppenstufen steigen?<br>1. Ja<br>2. Nein | a. Yes → Do you experience the shortage of breath as threatening?<br>1. Yes<br>2. No<br>b. No → Do you get out of breath when you walk more than 30 meters or climb 10 stairs?<br>1. Yes<br>2. No | a. Oui → Ressentez-vous le manque de soufflé menaçant?<br>1. Oui<br>2. Non<br>b. Non → Vous êtes essoufflé lorsque vous marchez sur plus de 30 mètres ou montez 10 marches?<br>1. Oui<br>2. Non |
| 16. Fühlen Sie sich schlapp oder abgeschlagen? | 16. Do you feel tired or worn out? | 16. Vous sentez-vous fatigué ou épuisé? |

| | | |
|---|---|---|
| a. Ja<br>b. Nein | a. Yes<br>b. No | a. Oui<br>b. Non |
| **Vorerkrankungen**<br>Risiko für die Möglichkeit eines schweren Covid-19 Verlaufs | **Previous diseases**<br>Risk of a severe Covid-19 course | **Maladies préexistantes**<br>Risque de possibilité d'un cours Covid-19 sévère |
| 17. Rauchen Sie oder haben Sie in den letzten 5 Jahren regelmäßig geraucht? | 17. Do you smoke or have you smoked regularly in the past 5 years? | 17. Fumez-vous ou avez-vous fumé régulièrement au cours des 5 dernières années? |
| a. Ja → Wie viele Zigaretten rauchen Sie durchschnittlich pro Tag?<br>1. < 5<br>2. 5-10<br>3. 10-20<br>4. > 20<br>b. Nein | a. Yes → How many cigarettes do you smoke per day on average?<br>1. < 5<br>2. 5-10<br>3. 10-20<br>4. > 20<br>b. No | a. Oui → Combien de cigarettes fumez-vous en moyenne par jour?<br>1. < 5<br>2. 5-10<br>3. 10-20<br>4. > 20<br>b. Non |
| 18. Haben Sie eine Lungenerkrankung (COPD, Asthma bronchiale, Lungenfibrose, Silikose) | 18. Do you have a lung disease (COPD, bronchial asthma, pulmonary fibrosis, silicosis) | 18. Avez-vous une maladie pulmonaire (MPOC, asthme bronchique, fibrose pulmonaire, silicose) |
| a. Ja → Welche Lungenerkrankung liegt bei Ihnen vor? (Mehrfachauswahl möglich)<br>1. COPD<br>2. Asthma<br>3. Lungenfibrose<br>4. Silikose<br>5. Sonstige, oben nicht benannt<br>b. Nein<br>c. Ich weiß es nicht | a. Yes → Which pulmonary disease do you have? (multiple answers are possible)<br>1. COPD<br>2. Asthma<br>3. Pulmonary fibrosis<br>4. Silicosis<br>5. Others, not mentioned above<br>b. No<br>c. I do not know | a. Oui → Quelle maladie pulmonaire avez-vous? (réponses multiples sont possibles)<br>1. MPOC<br>2. Asthme<br>3. Fibrose pulmonaire<br>4. Silicose<br>5. Autres<br>b. Non<br>c. Je ne sais pas |
| 19. Haben Sie Diabetes (Zuckerkrankheit)? | 19. Do you have diabetes? | 19. Avez-vous le diabète? |
| a. Ja<br>b. Nein<br>c. Ich weiß es nicht | a. Yes<br>b. No<br>c. I do not know | a. Oui<br>b. Non<br>c. Je ne sais pas |
| 20. Haben Sie eine Kreislauferkrankung? (Bluthochdruck, Herzschwäche, Stent, Herzklappenfehler) | 20. Do you have a circulatory disease? (High blood pressure, heart failure, stent, heart valve defects) | 20. Avez-vous une maladie circulatoire? (Hypertension artérielle, insuffisance cardiaque, stent, anomalies valvulaires cardiaques) |
| a. Ja<br>b. Nein<br>c. Ich weiß es nicht | a. Yes<br>b. No<br>c. I do not know | a. Oui<br>b. Non<br>c. Je ne sais pas |
| 21. Hatten Sie einen Schlaganfall oder eine Hirnblutung? | 21. Have you had a stroke or cerebral hemorrhage? | 21. Avez-vous eu une attaque cérébrale ou des hémorrhagies cérébrales? |
| a. Ja | a. Yes | a. Oui |

| | | |
|---|---|---|
| b. Nein<br>c. Ich weiß es nicht | b. No<br>c. I do not know | b. Non<br>c. Je ne sais pas |
| 22. Besteht bei Ihnen eine Tumorerkrankung? | 22. Do you have a cancer disease? | 22. Avez-vous une maladie de cancer? |
| a. Ja → Haben Sie eine Strahlentherapie und/oder Chemotherapie in den letzten 3 Monaten erhalten?<br>1. Ja<br>2. Nein<br>b. Nein<br>c. Ich weiß es nicht | a. Yes → Did you receive radiotherapy and/or chemotherapy during the last 3 months?<br>1. Yes<br>2. No<br>b. No<br>c. I do not know | a. Oui → Avez-vous reçu une radiothérapie et/ou une chimiothérapie au cours des 3 derniers mois?<br>1. Oui<br>2. Non<br>b. Non<br>c. Je ne sais pas |
| 23. Leiden Sie an einer chronischen entzündlichen Erkrankung (Rheuma, Morbus Crohn oder ähnliches)? | 23. Do you suffer from a chronic inflammatory disease (rheumatism, Crohn's disease or other)? | 23. Souffrez-vous d'une maladie inflammatoire chronique (rhumatismes, maladie de Crohn ou similaire)? |
| a. Ja → Nehmen Sie dauerhaft Cortison oder andere Immunsuppressiva ein? (Methotrexat, Cyclosporin, Biologicals, Mofetil etc.)<br>1. Ja<br>2. Nein<br>3. Ich weiß es nicht<br>b. Nein<br>c. Ich weiß es nicht | a. Yes → Do you regularly take cortisone or other immunosuppressants? (Methotrexate, cyclosporin, biologicals, mofetil etc.)<br>1. Yes<br>2. No<br>3. I do not know<br>b. No<br>c. I do not know | a. Oui → Prenez-vous de la cortisone ou d'autres immunosuppresseurs de façon permanente? (Méthotrexate, cyclosporine, produits biologiques, mofétil, etc.)<br>1. Oui<br>2. Non<br>3. Je ne sais pas<br>b. Non<br>c. Je ne sais pas |
| 24. Ist Ihre Nierenfunktion eingeschränkt? | 24. Is your kidney function impaired? | 24. Votre fonction rénale est-elle altérée? |
| a. Ja<br>b. Nein<br>c. Ich weiß es nicht | a. Yes<br>b. No<br>c. I do not know | a. Oui<br>b. Non<br>c. Je ne sais pas |
| 25. Ist bei Ihnen eine Allergie bekannt? (z.B. Pollen, Medikamente, Nahrungsmittel etc.) | 25. Do you have an allergy? (e.g. pollen, medication, food, etc.) | 25. Avez-vous une allergie? (p. ex. pollen, médicaments, nourriture, etc.) |
| a. Ja<br>b. Nein<br>c. Ich weiß es nicht | a. Yes<br>b. No<br>c. I do not know | a. Oui<br>b. Non<br>c. Je ne sais pas |
| 26. Haben Sie sich im Zeitraum von Oktober 2019 bis heute gegen Grippe impfen lassen? | 26. Have you been vaccinated against flu between October 2019 and today? | 26. Avez-vous été vacciné contre la grippe entre octobre 2019 et aujourd'hui? |
| a. Ja<br>b. Nein<br>c. Ich weiß es nicht | a. Yes<br>b. No<br>c. I do not know | a. Oui<br>b. Non<br>c. Je ne sais pas |
| 27. Sind Sie gegen Masern geimpft oder haben Sie eine Maserninfektion durchlebt? | 27. Have you been vaccinated against measles or have you had a measles infection? | 27. Avez-vous été vacciné contre la rougeole ou avez-vous eu une infection rougeoleuse? |

| | | |
|---|---|---|
| a. Ja<br>b. Nein<br>c. Ich weiß es nicht | a. Yes<br>b. No<br>c. I do not know | a. Oui<br>b. Non<br>c. Je ne sais pas |
| 28. Wie lautet Ihre Postleitzahl? | 28. What is your ZIP code? | 28. Quel est votre code postale? |
| FREITEXT | FREE TEXT | TEXTE LIBRE |
